# Association of the medical therapy with beta-blockers or inhibitors of renin-angiotensin system with clinical outcomes in patients with mildly reduced left ventricular ejection fraction after acute myocardial infarction

**DOI:** 10.1101/2022.03.01.22271686

**Authors:** Seung-Jae Joo, Song-Yi Kim, Jae-Geun Lee, Joon-Hyouk Choi, Hyeung Keun Park, Jong Wook Beom, Ki Yung Boo, Chang-Hwan Yoon, Jung-Hee Lee, Seung-Ho Hur, Jei Keon Chae, Myung Ho Jeong, the KAMIR-NIH registry investigators

## Abstract

In the era of the initial optimal interventional and medical therapy for acute myocardial infarction (AMI), a number of patients with mildly reduced left ventricular ejection fraction (EF) (41 - 49%) have been increasing. This observational study aimed to investigate the association between the medical therapy with oral beta-blockers or inhibitors of renin-angiotensin system (RAS) and 2-year clinical outcomes in patients with mildly reduced EF after AMI. Among patients enrolled in the Korea Acute Myocardial Infarction Registry-National Institute of Health, propensity-score matched patients who survived the initial attack and had mildly reduced EF were selected according to beta-blocker or RAS inhibitor therapy at discharge. Beta-blocker therapy at discharge was associated with lower 2-year major adverse cardiac events which was a composite of cardiac death, myocardial infarction, revascularization and re-hospitalization due to heart failure (8.7 vs. 12.8/100 patient-years; hazard ratio [HR] 0.68; 95% confidence interval [CI] 0.50-0.93; *P*=0.015), and no significant interaction between EF ≤45% and >45% was observed (*P*_interaction_=0.354). This association was mainly driven by lower myocardial infarction in patients with beta-blockers (HR 0.50; 95% CI 0.26-0.95; *P*=0.035). Inhibitors of RAS at discharge were associated with lower re-hospitalization due to heart failure (1.8 vs. 3.5/100 patient-years; HR 0.53; 95% CI 0.33-0.86; *P*=0.010) without a significant interaction between EF ≤45% and >45% (*P*_interaction_=0.333). In patients with mildly reduced EF after AMI, the medical therapy with beta-blockers or RAS inhibitors at discharge was associated with better 2-year clinical outcomes.

## Introduction

Heart failure (HF) had long been classified as HF with reduced ejection fraction (HFrEF) or HF with preserved EF (HFpEF). In most clinical trials, HFrEF was defined as EF less than 35-40%, and patients with EF above 40% were considered to have HFpEF. However, the 2016 European Society of Cardiology (ESC) guidelines for HF suggested an intermediate phenotype, i.e. HF with mid-range EF (EF between 40% to 49%) [1], and the 2021 ESC guidelines renamed it to HF with mildly reduced EF (HFmrEF) because retrospective analysis of clinical trials that included patients with EF in the 40 - 50% range showed some benefits from similar therapies to those with HFrEF [2].

An optimal medical therapy as well as early coronary reperfusion therapy is recommended in patients with acute myocardial infarction (AMI) to reduce cardiovascular mortality and morbidity. The evidence of clinical benefits of beta-blockers or inhibitors of renin-angiotensin system (RAS) after AMI is based on the clinical trials which enrolled patients with left ventricular (LV) EF below 40% or clinical HF [3-5]. Their effects in AMI patients with EF >40% have not been well documented. The clinical trials showing a long-term benefit of beta-blockers in patients with EF >40% after AMI are still lacking, and clinical studies of RAS inhibitors in patients with stable coronary artery disease (CAD) without LV systolic dysfunction or clinical HF showed inconsistent results [6-8].

In the era of early coronary reperfusion therapy, patients with mildly reduced EF after AMI have been increasing. However, the role of beta-blockers or RAS inhibitors in such patients is still unclear because they may be classified as having either preserved or reduced EF. We have already reported that beta-blocker therapy at discharge was associated with better 1-year clinical outcomes in patients with mildly reduced EF (>40%, <50%) as well as with reduced EF (≤40%) after AMI, but not in patients with preserved EF (≥50%) [9]. This finding suggested that AMI patients with mildly reduced EF be managed similarly to those with reduced EF. Nevertheless, because of different baseline characteristics of those patients with vs. without beta-blocker therapy at discharge, this association needed to be confirmed in the propensity score-matched patients. Also the role of RAS inhibitor therapy in AMI patients with mildly reduced EF was not analyzed in the previous study.

This observational study aimed to define the association between the medical therapy with beta-blockers or RAS inhibitors at discharge and 2-year clinical outcomes in patients with mildly reduced EF after AMI after propensity score matching.

## Methods

### Study population and data collection

The Korea Acute Myocardial Infarction Registry-National Institute of Health (KAMIR-NIH) is a nation-wide, prospective, observational, and on-line registry of South Korea from 20 university hospitals. Patients who were hospitalized primarily for AMI and signed informed consents were consecutively enrolled from Nov 2011 to Oct 2015. This study was conducted according to the Declaration of Helsinki. The study protocol was approved by the ethics committee at Chonnam National University Hospital, Republic of Korea (IRB No. CNUH-2011-172) and the institutional review boards of all participating hospitals approved the study protocol. Written informed consents were obtained from participating patients or legal representative. Data were collected by the attending physician with the assistance of a trained clinical research coordinator, via a web-based case report form in the clinical data management system of the Korea NIH [9, 10]. LV systolic function was evaluated with echocardiography during the initial hospitalization. Patients, who did not undergo echocardiographic study, died during index hospitalization, had incomplete clinical data or had EF ≤40% or ≥50%, were excluded.

AMI was diagnosed when there was an evidence of myocardial necrosis (a rise and/or fall in cardiac biomarker, preferably cardiac troponin), and at least one of the following: (1) symptoms of ischemia, (2) new or presumed new significant ST-segment-T wave changes or a new left bundle branch block, (3) a development of pathologic Q waves in the ECG, (4) an imaging evidence of the new loss of viable myocardium or new regional wall motion abnormality, and (5) the identification of an intracoronary thrombus by angiography [11]. Coronary reperfusion included reperfusion by percutaneous coronary intervention (PCI), thrombolysis, or coronary artery bypass graft (CABG), myocardial infarction with non-obstructed coronary arteries (MINOCA) [12], and myocardial bridge. RAS inhibitors included angiotensin-converting enzyme inhibitors (ACEi) and angiotensin receptor blockers (ARB).

### Clinical endpoints and definition

The primary endpoint was 2-year major adverse cardiac events (MACE) which was a composite of cardiac death, myocardial infarction (MI), revascularization, and re-hospitalization due to HF. The secondary endpoints were each component of MACE, all-cause death, stroke, 2-year major adverse cardiac and cerebrovascular events (MACCE) which was a composite of the primary endpoint and stroke, and 2-year MACE with non-cardiac death.

All deaths were considered to be associated with cardiac problems, unless a definite non-cardiac cause was established. MI included re-infarction or recurrent MI [11]. Revascularization included repeated PCI or CABG on either target or non-target vessels. The staged PCI was excluded from revascularization. The clinical follow-ups were routinely performed at 6-, 12-, 24-, and 36-month by visiting the hospital or whenever any clinical events occurred. If patients did not visit the hospitals, the outcome data were assessed by telephone interview. Clinical events were not centrally adjudicated. The patient’s physician identified all events and the principal investigator of each hospital confirmed them.

### Statistical analysis

Because either beta-blocker or RAS inhibitor therapy was not randomized, propensity score matching was performed as a sensitivity analysis to minimize selection or predisposition bias. The propensity score was estimated using multiple logistic regression analysis with all variables in Table 1 and 2. Using a greedy nearest matching algorithm with 0.1 caliper width, each patient without beta-blockers or RAS inhibitors was matched to a maximum of two patients in beta-blocker or RAS inhibitor group. The efficacy of the propensity score model was assessed by estimating standardized differences for each covariate between groups.

**Table 1.** Baseline characteristics of patients with vs. without beta-blockers at discharge after propensity-score matching.

|  | All patients<br>(N=1,048) | With BB<br>(N=685) | Without BB<br>(N=363) | SD | *P* value |
| --- | --- | --- | --- | --- | --- |
| Age (years) | 66.6±11.8 | 66.4±11.3 | 66.8±12.7 | <0.001 | 0.635 |
| Male | 736 (70.2) | 480 (70.1) | 256 (70.5) | 0.027 | 0.887 |
| Body mass index (kg/m<sup>2</sup>) | 23.42±3.21 | 23.50±3.24 | 23.27±3.17 | -0.044 | 0.266 |
| Hypertension | 502 (47.9) | 325 (47.4) | 177 (48.8) | 0.025 | 0.697 |
| Diabetes mellitus | 284 (27.1) | 186 (27.2) | 98 (27.0) | 0.006 | 1.000 |
| Prior angina pectoris | 105 (10.0) | 65 (9.5) | 40 (11.0) | 0.035 | 0.450 |
| Prior myocardial infarction | 94 (9.0) | 63 (9.2) | 31 (8.5) | -0.014 | 0.820 |
| Prior heart failure | 9 (0.9) | 6 (0.9) | 3 (0.8) | <0.001 | 1.000 |
| Prior stroke | 74 (7.1) | 47 (6.9) | 27 (7.4) | 0.021 | 0.800 |
| Current smoker | 397 (37.9) | 262 (38.2) | 135 (37.2) | -0.014 | 0.789 |
| Killip class ≥II | 262 (25.0) | 165 (24.1) | 97 (26.7) | 0.040 | 0.369 |
| eGFR <60 mL/min/1.73m<sup>2</sup> | 228 (21.8) | 149 (21.8) | 79 (21.8) | -0.010 | 1.000 |
| Left ventricular EF (%) | 45.7±2.5 | 45.7±2.4 | 45.5±2.5 | -0.072 | 0.282 |
| STEMI | 546 (52.1) | 354 (51.7) | 192 (52.9) | 0.041 | 0.745 |
| Coronary reperfusion<sup>a</sup> | 1,005 (95.9) | 656 (95.8) | 349 (96.1) | 0.023 | 0.871 |
| Medications at discharge |  |  |  |  |  |
| Aspirin | 1,047 (99.9) | 685 (100) | 362 (99.7) | -0.038 | 0.346 |
| P2Y12 inhibitor | 1,022 (95.6) | 655 (95.6) | 347 (95.6) | 0.015 | 1.000 |
| RAS inhibitors | 627 (59.8) | 420 (61.3) | 207 (57.0) | -0.019 | 0.186 |
| Statins | 947 (90.4) | 619 (90.4) | 328 (90.4) | 0.041 | 1.000 |
| Values are mean ± standard deviation or number (%).<br>Abbreviations: BB, beta-blockers; EF, ejection fraction; eGFR, estimated glomerular filtration rate by MDRD equation; RAS, renin-angiotensin system; SD, standardized difference; STEMI, ST elevation myocardial infarction.<br><sup>a</sup>Includes reperfusion by percutaneous coronary intervention, thrombolysis, coronary artery bypass graft, or myocardial infarction with non-obstructed coronary arteries (MINOCA). |  |  |  |  |  |

**Table 2.** Baseline characteristics of patients with vs. without inhibitors of renin-angiotensin system at discharge after propensity-score matching.

|  | All patients<br>(N=1,559) | With RASI<br>(N=1,003) | Without RASI<br>(N=556) | SD | *P* value |
| --- | --- | --- | --- | --- | --- |
| Age (years) | 65.2±12.5 | 65.0±12.4 | 65.5±12.6 | 0.002 | 0.429 |
| Male | 1,129 (72.4) | 723 (72.1) | 406 (73.0) | 0.032 | 0.723 |
| Body mass index (kg/m<sup>2</sup>) | 23.42±3.07 | 23.42±3.01 | 23.43±3.17 | 0.033 | 0.960 |
| Hypertension | 712 (45.7) | 457 (45.6) | 255 (45.9) | 0.007 | 0.916 |
| Diabetes mellitus | 460 (29.5) | 292 (29.1) | 168 (30.2) | 0.017 | 0.643 |
| Prior angina pectoris | 143 (9.2) | 84 (8.4) | 59 (10.6) | 0.052 | 0.144 |
| Prior myocardial infarction | 139 (8.9) | 90 (9.0) | 49 (8.8) | <0.001 | 1.000 |
| Prior heart failure | 21 (1.3) | 14 (1.4) | 7 (1.3) | -0.025 | 1.000 |
| Prior stroke | 113 (7.2) | 72 (7.2) | 41 (7.4) | -0.010 | 0.919 |
| Current smoker | 605 (38.8) | 392 (39.1) | 213 (38.3) | 0.004 | 0.786 |
| Killip class ≥II | 358 (23.0) | 230 (22.9) | 128 (23.0) | -0.011 | 1.000 |
| eGFR <60 mL/min/1.73m<sup>2</sup> | 329 (21.1) | 206 (20.5) | 123 (22.1) | 0.009 | 0.476 |
| Left ventricular EF (%) | 45.5±2.5 | 45.5±2.5 | 45.6±2.5 | 0.025 | 0.613 |
| STEMI | 920 (59.0) | 603 (60.1) | 317 (57.0) | -0.031 | 0.237 |
| Coronary reperfusion<sup>a</sup> | 1,504 (96.5) | 969 (96.6) | 535 (96.2) | <0.001 | 0.671 |
| Medications at discharge |  |  |  |  |  |
| Aspirin | 1,556 (99.8) | 1,001 (99.8) | 555 (99.8) | 0.015 | 1.000 |
| P2Y12 inhibitor | 1,505 (96.5) | 969 (96.6) | 536 (96.4) | 0.016 | 0.885 |
| Beta-blockers | 1,199 (76.9) | 800 (79.8) | 399 (71.8) | -0.008 | <0.001 |
| Statins | 1,439 (92.3) | 931 (92.8) | 508 (91.4) | -0.015 | 0.322 |
| Values are mean ± standard deviation or number (%).<br>Abbreviations: EF, ejection fraction; eGFR, estimated glomerular filtration rate by MDRD equation; RASI, inhibitors of renin-angiotensin system; SD, standardized difference; STEMI, ST elevation myocardial infarction.<br><sup>a</sup>Includes reperfusion by percutaneous coronary intervention, thrombolysis, coronary artery bypass graft, myocardial bridge, or myocardial infarction with non-obstructed coronary arteries (MINOCA). |  |  |  |  |  |

Data were expressed as mean ± standard deviation or median (interquartile range) for continuous variables, and as number (percentage) for categorical variables. Data were compared using unpaired t-test for continuous variables, and chi-square test for categorical variables. Survival curves for clinical endpoints and cumulative event rates with incidence rates per 100 patient-years were generated using Kaplan–Meier estimates. Hazard ratios (HR) and their 95% confidence interval (CI) for each clinical endpoint were calculated using Cox proportional hazard analysis. In multivariate Cox regression analysis, age, sex, body mass index (BMI), hypertension, diabetes mellitus (DM), prior angina, prior MI, prior HF, current smoker, Killip class, estimated glomerular filtration rate (eGFR) by Modification of Diet in Renal Disease (MDRD) equation, LVEF, type of MI, coronary reperfusion, and medications (aspirin, P2Y12 inhibitors, beta-blockers or RAS inhibitors, and statins) at discharge were included as covariates. Subgroups that were defined post-hoc according to demographic and clinical characteristics included age (<65, ≥65-<80, & ≥80 years), sex, hypertension, DM, prior MI, current smoker, Killip class ≥2, eGFR<60 mL/min/1.73m^2^, EF (≤45% & >45%), ST-elevation myocardial infarction (STEMI), and beta-blockers or RAS inhibitors at discharge.

All statistical analyses were performed with the statistical package SPSS version 23 (IBM Co, Armonk, NY, US) and R version 3.1.3 (R Foundation for Statistical Computing,Vienna, Austria). Clinical significance was defined as *P* <0.05.

## Results

Total 13,624 consecutive patients were enrolled in the KAMIR-NIH. After excluding 10,720 patients (1,153 patients without echocardiographic data, 252 patients who died during index hospitalization, 19 patients with incomplete data, 1,670 patients with EF ≤40%, and 7,626 patients with EF ≥50%), 2,904 patients with EF >40%, <50% were analyzed in this study. They comprised 24% of patients who underwent echocardiographic study and survived the initial attack. Beta-blockers or RAS inhibitors were prescribed at the discretion of attending physicians. They were taken in 2,508 (86%) and 2,316 patients (80%) at discharge, respectively. After propensity score matching, 1,048 patients with or without beta-blockers at discharge and 1,559 patients with or without RAS inhibitors at discharge were selected (Fig 1). Overall reperfusion rate was 96%, and PCI with drug-eluting stents was the main method of coronary reperfusion in both propensity-score matched cohorts (S1 and S2 Table).

**Fig 1.**
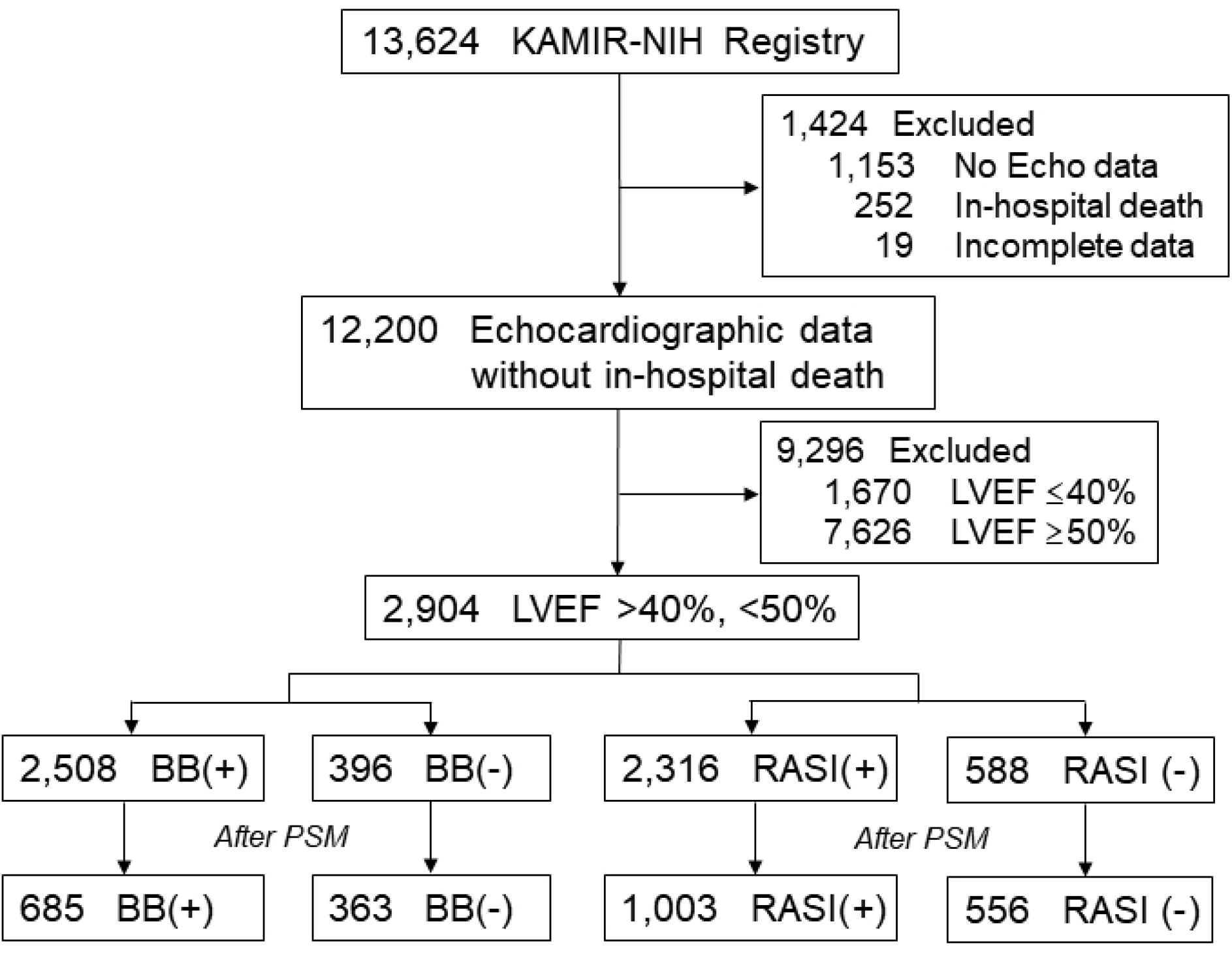
Selection of patients for analysis. BB, beta-blockers; Echo, echocardiography; KAMIR-NIH, Korean Acute Myocardial Infarction Registry-National Institute of Health; LVEF, left ventricular ejection fraction, PSM, propensity score-matching; RASI, inhibitors of renin-angiotensin system.

In the entire cohort, patients without beta-blockers at discharge were older, and had lower BMI, more prior angina, more Killip class ≥II, more eGFR <60 mL/min/1.73m^2^, less STEMI, and less coronary reperfusion. Less P2Y12 inhibitors, RAS inhibitors, or statins were prescribed at discharge (S3 Table). After propensity score matching, baseline characteristics of 685 patients with beta-blockers and 363 patients without beta-blockers at discharge were well balanced (Table 1). Likewise, patients without RAS inhibitors in the entire cohort were older, and had lower BMI, more prior angina, more eGFR <60 mL/min/1.73m^2^, less STEMI, and less coronary reperfusion. They were taking less P2Y12 inhibitors, beta-blockers, or statins at discharge (S4 Table). After propensity score matching, baseline characteristics of 1,003 patients with RAS inhibitors and 556 patients without RAS inhibitors at discharge were well balanced (Table 2).

In propensity-score matched cohorts, 2-year follow-up rates were 96% and 92% in patients with and without beta-blockers at discharge, and 95% and 94% in patients with and without RAS inhibitors at discharge, respectively. Beta-blockers were discontinued at 1- and 2-year in 9% and 18% of survived patients with beta-blockers at discharge, but were taken at 1- and 2-year in 45% and 41% of patients without them at discharge, respectively. RAS inhibitors were stopped at 1- and 2-year in 19% and 20% of survived patients with RAS inhibitors at discharge, but were taken at 1- and 2-year in 37% and 46% of patients without them at discharge, respectively.

Patients with beta-blockers at discharge showed significantly lower 2-year MACE (8.7 vs. 12.8/100 patient-years) and HR was 0.68 (95% CI; 0.50-0.93; *P*=0.015) after full adjustment, and this association was mainly driven by lower MI (HR 0.50; 95% CI; 0.26-0.95; *P*=0.035) and revascularization (HR 0.62; 95% CI; 0.41-0.95; *P*=0.030) (Table 3, Fig 2). Likewise, 1-year MACE, cardiac deaths, and MI were significantly lower in patients with beta-blockers (S5 Table). The association between beta-blocker therapy at discharge and 2-year MACE appeared to be consistent across a series of subgroups (Fig 3). No significant interaction between EF ≤45% and >45% in terms of any 2-year clinical endpoints was noted (S1 Fig). Beta-blocker therapy at discharge was also associated with lower 2-year MACE with non-cardiac death (Table 3). Carvedilol (50%), bisoprolol (43%) and nebivolol (5%) were the major beta-blockers that prescribed at discharge (S6 Table). All beta-blockers were used in lower doses than those recommended in the guidelines. They showed comparable clinical outcomes without a significant interaction (S2 Fig).

**Table 3.** Multivariate Cox-proportional hazard analysis of 2-year events in patients with vs. without beta-blockers at discharge after propensity-score matching.

| Events | With BB<br>(N=685) | Without BB<br>(N=363) | Hazard ratio<sup>a</sup><br>(95% CI) | *P* value |
| --- | --- | --- | --- | --- |
| MACE | 103 (8.7) | 73 (12.8) | 0.68 (0.50-0.93) | 0.015 |
| Cardiac death | 32 (2.5) | 25 (4.0) | 0.67 (0.38-1.16) | 0.154 |
| All-cause death | 54 (4.3) | 38 (6.1) | 0.69 (0.45-1.05) | 0.085 |
| Myocardial infarction | 20 (1.6) | 19 (3.1) | 0.50 (0.26-0.95) | 0.035 |
| Revascularization | 50 (4.1) | 38 (6.5) | 0.62 (0.41-0.95) | 0.030 |
| Heart failure<sup>b</sup> | 29 (2.4) | 14 (2.3) | 1.06 (0.55-2.03) | 0.868 |
| Stroke | 12 (1.0) | 9 (1.4) | 0.69 (0.29-1.66) | 0.409 |
| MACCE | 111 (9.5) | 78 (13.7) | 0.70 (0.52-0.94) | 0.017 |
| MACE with NCD | 124 (10.5) | 82 (14.3) | 0.72 (0.54-0.96) | 0.025 |
| Values are number of patients with events (rate per 100 patient-years)<br>Abbreviations: BB, beta-blockers, CI, confidence interval; MACCE, major adverse cardiac and cerebrovascular event; MACE, major adverse cardiac event; NCD, Non-cardiac death<br><sup>a</sup>Adjusted for age, sex, body mass index, hypertension, diabetes mellitus, prior angina, prior MI, prior heart failure, current smoker, Killip class, estimated glomerular filtration rate, left ventricular ejection fraction, type of myocardial infarction, coronary reperfusion, and medications (aspirin, P2Y12 inhibitors, inhibitors of renin-angiotensin system and statins) at discharge<br><sup>b</sup>Re-hospitalization due to heart failure |  |  |  |  |

**Fig 2.**
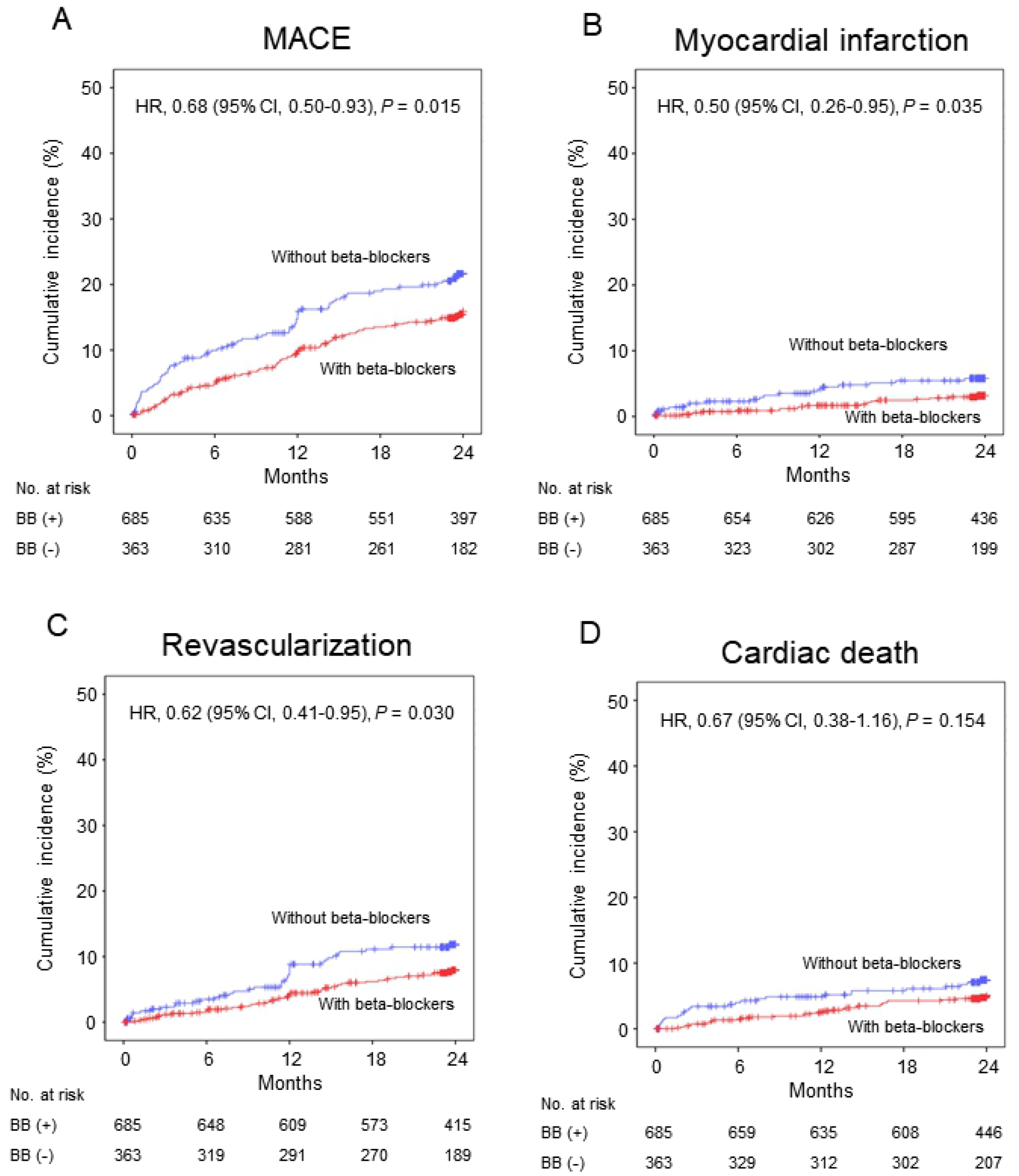
Kaplan-Meier curves and adjusted hazard ratios with 95% confidence intervals for 2-year events with vs. without beta-blockers at discharge in the propensity score-matched cohorts. (A) major adverse cardiac events (B) myocardial infarction (C) revascularization (D) cardiac death. BB, beta-blockers; CI, confidence interval; HR hazard ratio; MACE, major adverse cardiac events.

**Fig 3.**
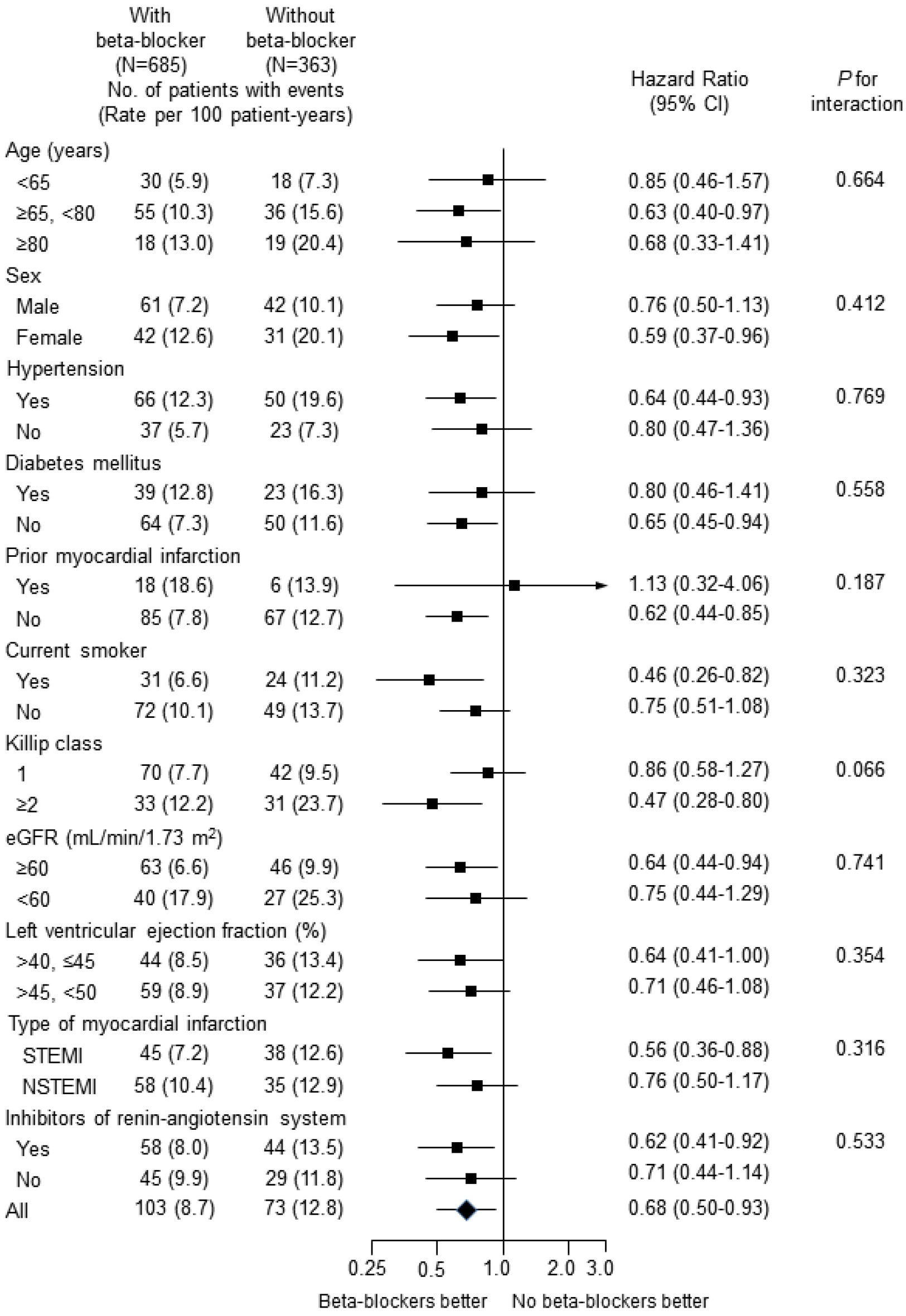
Adjusted hazard ratios of 2-year major adverse cardiac events for subgroups in the propensity score-matched cohort with vs. without beta-blockers at discharge. CI, confidence interval; eGFR, estimated glomerular filtration rate by MDRD equation; NSTEMI, non-ST elevation myocardial infarction; STEMI, ST-elevation myocardial infarction.

However, no significant association of RAS inhibitors at discharge with 2-year clinical endpoints except re-hospitalization due to HF was observed after full adjustment (Table 4, Fig 4). Patients with RAS inhibitors at discharge were associated with lower 2-year re-hospitalization due to HF (1.8 vs. 3.5/100 patient-years; HR 0.53; 95% CI; 0.33-0.86; *P*=0.010). Likewise, 1-year re-hospitalization due to HF was significantly lower in patients with RAS inhibitors at discharge (S7 Table). The association between RAS inhibitor therapy at discharge and 2-year re-hospitalization due to HF appeared to be consistent across a series of subgroups (S3 Fig), and no significant interaction between EF ≤45% and >45% was shown (S4 Fig). Perindopril (50%) and ramipril (40%) were the major ACEi’s, and candesartan (35%), losartan (24%), telmisartan (20%) and valsartan (14%) were the major ARB’s that prescribed at discharge (S8 Table). All RAS inhibitors were used in lower doses than those recommended in the guidelines. ACEi and ARB showed comparable clinical outcomes without a significant interaction (Fig 5).

**Table 4.** Multivariate Cox-proportional hazard analysis of 2-year events patients with vs. without inhibitors of renin-angiotensin system at discharge after propensity-score matching.

| Events | With RASI<br>(N=1,003) | Without RASI<br>(N=556) | Hazard ratio<sup>a</sup><br>(95% CI) | *P* value |
| --- | --- | --- | --- | --- |
| MACE | 151 (8.9) | 89 (9.6) | 0.95 (0.73-1.23) | 0.676 |
| Cardiac death | 42 (2.3) | 27 (2.7) | 0.91 (0.55-1.49) | 0.700 |
| All-cause death | 75 (4.1) | 45 (4.5) | 0.96 (0.66-1.39) | 0.809 |
| Myocardial infarction | 28 (1.6) | 18 (1.8) | 0.91 (0.50-1.67) | 0.763 |
| Revascularization | 83 (4.8) | 39 (4.0) | 1.22 (0.83-1.79) | 0.306 |
| Heart failure<sup>b</sup> | 33 (1.8) | 34 (3.5) | 0.53 (0.33-0.86) | 0.010 |
| Stroke | 15 (0.8) | 12 (1.2) | 0.72 (0.33-1.55) | 0.395 |
| MACCE | 163 (9.7) | 98 (10.6) | 0.92 (0.71-1.18) | 0.514 |
| MACE with NCD | 182 (10.7) | 102 (11.0) | 0.99 (0.78-1.27) | 0.954 |
| Values are number of patients with events (rate per 100 patient-years)<br>Abbreviations: CI, confidence interval; MACCE, major adverse cardiac and cerebrovascular event; MACE, major adverse cardiac event; NCD, Non-cardiac death; RASI, inhibitors of renin-angiotensin system.<br><sup>a</sup>Adjusted for age, sex, body mass index, hypertension, diabetes mellitus, prior angina, prior MI, prior heart failure, current smoker, Killip class, estimated glomerular filtration rate, left ventricular ejection fraction, type of myocardial infarction, coronary reperfusion, and medications (aspirin, P2Y12 inhibitors, beta-blockers and statins) at discharge<br><sup>b</sup>Re-hospitalization due to heart failure |  |  |  |  |

**Fig 4.**
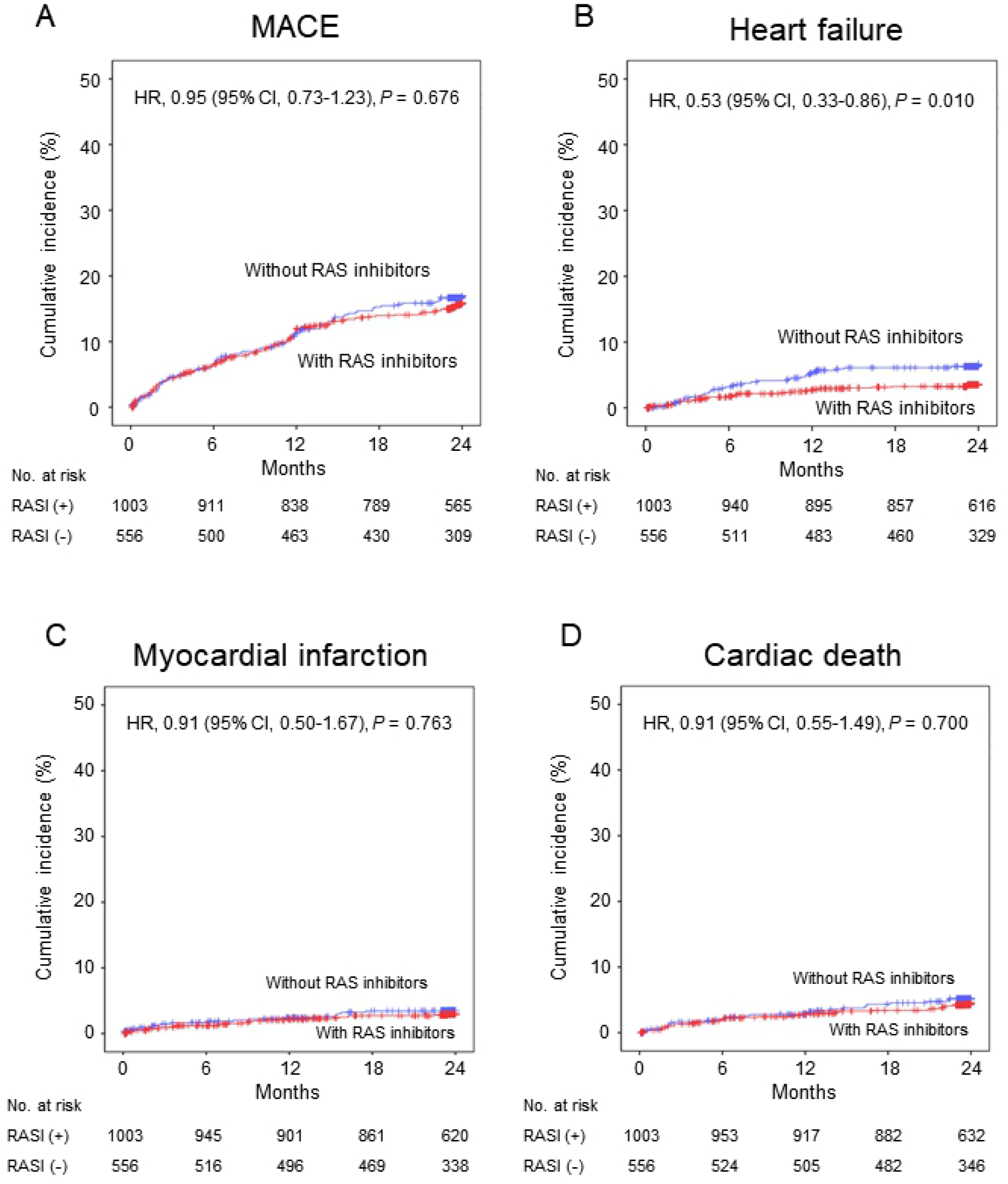
Kaplan-Meier curves and adjusted hazard ratios with 95% confidence intervals for 2-year events with vs. without inhibitors of renin-angiotensin system at discharge in the propensity score-matched cohorts. (A) major adverse cardiac events (B) heart failure (C) myocardial infarction (D) cardiac death. CI, confidence interval; HR hazard ratio; MACE, major adverse cardiac events; RASI, inhibitors of renin-angiotensin system.

**Fig 5.**
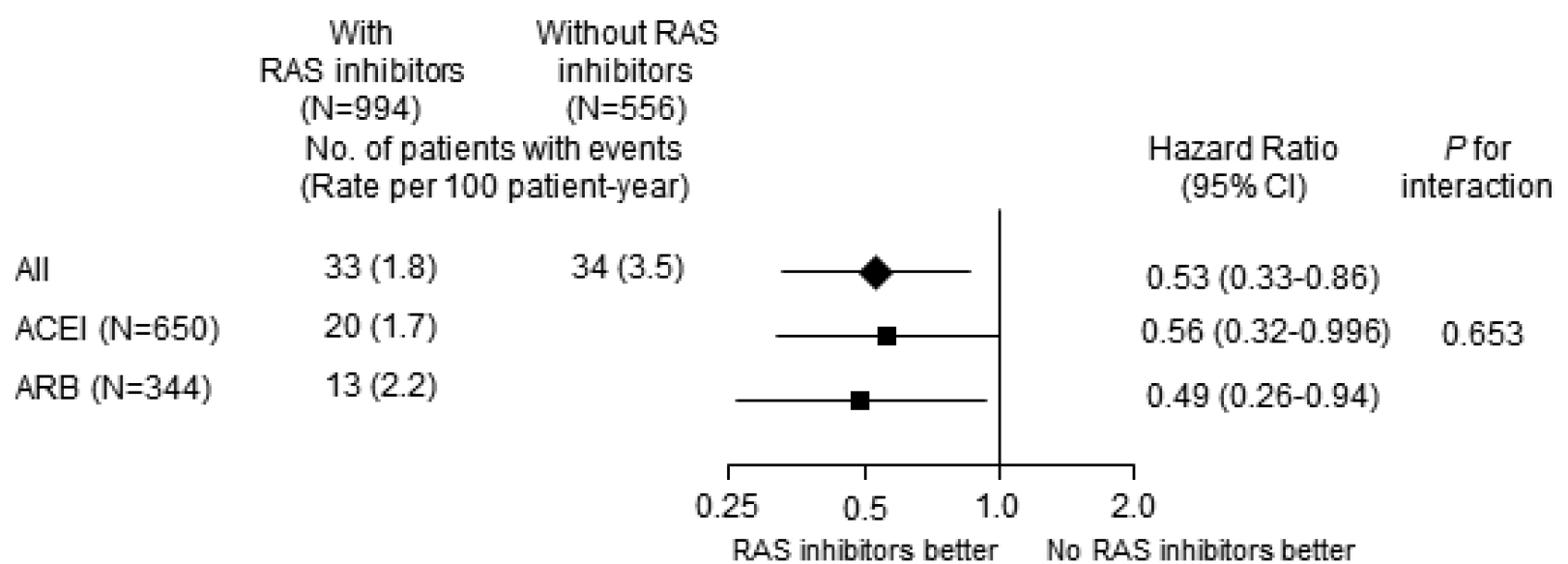
Adjusted hazard ratios of 2-year re-hospitalization due to heart failure for angiotensin-converting enzyme inhibitors vs. angiotensin receptor blockers at discharge in the propensity score-matched cohort. ACEI, angiotensin-converting enzyme inhibitors; ARB, angiotensin receptor blockers; CI, confidence interval.

## Discussion

The main finding of this study is that the medical therapy with beta-blockers or RAS inhibitors at discharge was associated with better 2-year clinical outcomes without significant interaction between EF ≤45% and >45% in patients with mildly reduced EF after AMI in the era of early coronary reperfusion and the contemporary other optimal medical therapy with antiplatelet agents and statins.

The prevalence of HFmrEF in the registries and clinical trials of HF patients was 14-23% [13]. Similarly to HFrEF, the most common etiology of HFmrEF was an ischemic heart disease (IHD) [14], and prior MI was an important predictor of HFmrEF [15]. Our data showed that a quarter of patients with AMI had mildly reduced EF. All-cause mortality of patients with HFmrEF was intermediate between HFrEF and HFpEF [14, 15], and CAD indicated a higher risk of death in HFmrEF [16]. Therefore, patients with mildly reduced EF as well as reduced EF after AMI should be managed with an optimal medical therapy to reduce cardiovascular mortality and morbidity.

Beta-blockers’ benefit in patients with HFrEF was clearly shown in previous clinical trials, and evidence-proven beta-blockers are strongly recommended in the guidelines [1, 2, 17, 18]. However, the role of beta-blockers in patients with HFpEF or HFmrEF was not clearly proven except one meta-analysis of clinical trials which showed 41% and 52% reduction of all-cause and cardiac mortality in patients with HFmrEF in sinus rhythm [19]. But the median EF of 40% (interquartile range 40-43%) in this meta-analysis could not provide the sufficient evidence to support the use of beta-blockers in patients with HFmrEF, especially when EF is >45%. In HF registries, beta-blocker therapy was associated with lower all-cause mortality in patients with HFmrEF [16, 20].

Likewise, no randomized clinical trials of beta-blockers in AMI patients with mildly reduced EF are available so far. In this regard, registry data may provide evidence about an optimal medical therapy in these patients despite the inherent limitations. Previous registry data showed no association of beta-blocker therapy with reduced mortality among survivors of AMI without HF or LV systolic dysfunction (EF <30-40%) [21, 22]. In our registry, beta-blocker therapy at discharge was associated with lower 2-year MACE in AMI patients with mildly reduced EF without significant interaction between EF ≤45% and >45%. This association was mainly driven by lower MI, and revascularization which was partially associated with recurrent MI. AMI patients with mildly reduced EF have scarred or non-viable myocardium to some extent, and in this clinical setting, beta-blocker therapy may be effective in reducing fatal arrhythmia, myocardial ischemia, or recurrent MI [9, 22]. Our data suggest that beta-blocker therapy be considered in patients with mildly reduced EF after AMI.

In MI patients with reduced EF or clinical HF, ACEi therapy was shown to reduce rates of death, MI and re-admission for HF [4], and ARB was non-inferior to ACEi with regard to all-cause mortality and combined end point of cardiovascular death, recurrent MI or hospitalization for HF [5]. In patients with HFmrEF, a registry data showed RAS inhibitors were associated with lower all-cause mortality [16], and in re-analyzed data of the Candesartan in Heart Failure-Assessment of Reduction in Mortality and Morbidity (CHARM) Programme, candesartan significantly reduced hospitalization for HF in HFmrEF, but not in HFpEF [23].

In patients with stable CAD without LV systolic dysfunction (EF <40%) or clinical HF, two large clinical trials showed ACEi’s role in reducing cardiovascular death and MI [6, 7], but other trial failed to provide further clinical benefit [8]. ARB in patients with stable CAD also did not reduce cardiac death, MI or hospitalization for HF [24]. This inconsistent result may be caused by the different characteristics of enrolled patients and the rates of clinical events which were affected by the intensive risk modulating medical therapy such as anti-platelet agents, beta-blockers or statins, and optimal coronary revascularization [8]. In a meta-analysis of clinical trials in stable CAD without HF, RAS inhibitors lowered all-cause mortality, cardiovascular mortality and HF when compared with placebo, but their benefit on all-cause and cardiovascular mortality was shown only in trials with high control event rates [25]. However, the role of RAS inhibitors in AMI patients with mildly reduced EF has not been proven yet in randomized clinical trials. Only a registry data of HFmrEF showed the mortality benefit of RAS inhibitor therapy in patients with CAD [16]. In our registry, RAS inhibitor therapy at discharge was associated with lower 2-year re-hospitalization due to HF in AMI patients with mildly reduced EF without significant interaction between EF ≤45% and >45% despite early coronary reperfusion and other guideline-directed medical therapy, and this outcome was comparable between ACEi and ARB. HFmrEF may be in the transition stage that becomes to improve to HFpEF or to progress to HFrEF, and IHD etiology was associated with a decrease in EF [20]. The activation of RAS after AMI causes LV dilatation and dysfunction which may induce clinical HF, and this remodeling process was shown to be attenuated by RAS inhibition.[4, 5] Our data suggest that RAS inhibitor therapy be considered in AMI patients with mildly reduced EF to prevent HF hospitalization.

### Limitations

First, this study analyzed a non-randomized, observational registry data. Beta-blockers or RAS inhibitors were prescribed at the discretion of an attending physician. The information why physicians did not prescribe beta-blockers or RAS inhibitors to some patients at discharge was not available. Although we tried to adjust the potential confounding factors by multivariable and a propensity score-matched analysis, other unrecorded or residual confounders as well as selection bias could not be completely controlled. Therefore, our results should be interpreted with caution. Second, because patients’ medications were recorded only at discharge, 1-year and 2-year, we could not ascertain whether patients actually obtained them, took them as prescribed, and adhered for two years. In addition, a large cross-over was observed in patients without beta-blockers or RAS inhibitors at discharge, and 41% and 46% of those patients were taking beta-blockers and RAS inhibitors at 2-year, respectively. However, despite this cross-over, taking beta-blockers or RAS inhibitors from the hospital discharge was associated with improved clinical outcomes. Third, beta-blockers and RAS inhibitors at discharge were prescribed at only a quarter to half of maximal dose recommended in the guidelines, and the individual dose at the time of clinical events was not available. However, in an American registry of AMI, beta-blockers were prescribed less than a quarter of maximal dose at discharge in 60% of patients, and the lowest mortality was observed in >12.5%-25% dose group [26]. In the Swedish Heart Failure Registry, RAS inhibitors of >50% of target dose, compared with ≤50% of target dose, showed better mortality benefit in patients with HFpEF (EF ≥40%), but over 70% of patients were prescribed ≤50% of target dose [27]. The lower than maximal recommended dose may be a usual prescription pattern in “real-world” registries, and the optimal dose of beta-blockers or RAS inhibitors in patients with AMI with mildly reduced EF needs to be confirmed in a randomized clinical trial.

## Conclusions

In patients with mildly reduced EF after AMI, the medical therapy with beta-blockers or RAS inhibitors at discharge were associated with better 2-year clinical outcomes without significant interaction between EF ≤45% and >45%. These data suggest that long-term beta-blocker or RAS inhibitor therapy need to be considered in AMI patients with mildly reduced EF.

## Data Availability

All relevant data are within the manuscript and its Supporting Information files. The original raw data cannot be shared publicly in order to protect patient confidentiality. The raw data are available from the Jeju National University University Hospital Institutional Data Access / Ethics Committee (contact via) to researchers who meet the criteria for access to confidential data.

## Acknowledgements

We appreciate the contribution of other KAMIR-NIH investigators: Tae Hoon Ahn, Ki-Bae Seung, Chong-Jin Kim, Shung Chull Chae, Jin-Yong Hwang, Seung-Woon Rha, Kwang Soo Cha, Hyo-Soo Kim, Hyeon-Cheol Gwon, Seok Kyu Oh, Junghan Yoon, In-Whan Seong, Kyung-Kuk Hwang, Doo-Il Kim

## Supporting information

**S1 Table. Reperfusion rates and methods in patients with vs. without beta-blockers at discharge after propensity score matching**

**S2 Table. Reperfusion rates and methods in patients with vs. without inhibitors of renin-angiotensin system at discharge after propensity score matching**

**S3 Table. Baseline characteristics of patients with vs. without beta-blockers at discharge in the entire cohort**

**S4 Table. Baseline characteristics of patients with vs. without inhibitors of renin-angiotensin system at discharge in the entire cohort**

**S5 Table. Multivariate Cox-proportional hazard analysis of 1-year events in patients with vs. without beta-blockers at discharge after propensity score matching**

**S6 Table. Generic names and doses of beta-blockers that prescribed at discharge in propensity-score matched patients**

**S7 Table. Multivariate Cox-proportional hazard analysis of 1-year events in patients with vs. without inhibitors of renin-angiotensin system at discharge after propensity score matching**

**S8 Table. Generic names and doses of inhibitors of renin-angiotensin system that prescribed at discharge in propensity-score matched patients**

**S1 Fig. Adjusted hazard ratios of 2-year events according to left ventricular ejection fraction ≤45% vs. >45% in the propensity score-matched cohort with or without beta-blockers at discharge**. CI, confidence interval; LVEF, left ventricular ejection fraction; MACCE, major adverse cardiac and cerebrovascular event; MACE, major adverse cardiac event.

**S2 Fig. Adjusted hazard ratios of 2-year major adverse cardiac events in the propensity score-matched cohort according to generic names of beta-blockers at discharge**. (A) adjusted hazard ratios with vs. without beta-blockers. (B) adjusted hazard ratios among beta-blockers. CI, confidence interval.

**S3 Fig. Adjusted hazard ratios of 2-year re-hospitalization due to heart failure for subgroups in the propensity score-matched cohort with vs. without inhibitors of renin-angiotensin system at discharge**. CI, confidence interval; eGFR, estimated glomerular filtration rate by MDRD equation; NSTEMI, non-ST elevation myocardial infarction; STEMI, ST-elevation myocardial infarction.

**S4 Fig. Adjusted hazard ratios of 2-year events according to left ventricular ejection fraction ≤45% vs. >45% in the propensity score-matched cohort with or without inhibitors of renin-angiotensin system at discharge**. CI, confidence interval; LVEF, left ventricular ejection fraction; MACCE, major adverse cardiac and cerebrovascular event; MACE, major adverse cardiac event.

## Author Contributions

**Conceptualization:** Seung-Jae Joo, Hyeung Keun Park, Chang-Hwan Yoon, Jung-Hee Lee, Seung-Ho Hur, Jei Keon Chae, Myung Ho Jeong

**Data curation:** Seung-Jae Joo, Song-Yi Kim, Jae-Geun Lee, Joon-Hyouk Choi, Jong Wook Beom, Ki Yung Boo

**Formal analysis:** Seung-Jae Joo, Joon-Hyouk Choi. Hyeung Keun Park.

**Funding acquisition:** Myung Ho Jeong.

**Investigation:** Seung-Jae Joo, Chang-Hwan Yoon, Jung-Hee Lee, Seung-Ho Hur, Jei Keon Chae, Myung Ho Jeong

**Methodology:** Seung-Jae Joo, Hyeung Keun Park

**Project administration:** Seung-Jae Joo, Myung Ho Jeong

**Resources:** Seung-Jae Joo, Myung Ho Jeong.

**Software:** Seung-Jae Joo, Joon-Hyouk Choi. Hyeung Keun Park

**Supervision:** Seung-Jae Joo.

**Validation:** Hyeung Keun Park.

**Visualization:** Seung-Jae Joo.

**Writing – original draft:** Seung-Jae Joo.

**Writing – review & editing: :** Seung-Jae Joo, Song-Yi Kim, Jae-Geun Lee, Joon-Hyouk Choi, Jong Wook Beom, Ki Yung Boo,. Hyeung Keun Park, Chang-Hwan Yoon, Jung-Hee Lee, Seung-Ho Hur, Jei Keon Chae, Myung Ho Jeong

## Notes

### Competing Interest Statement

The authors have declared no competing interest.

### Funding Statement

MJ received a fund [2016-ER6304-02] by Korea Centers for Disease Control and Prevention. The funders had no role in study design, data collection and analysis, decision to publish, or preparation of the manuscript.

### Author Declarations

The ethics committee at Chonnam National University Hospital (CNUH-2011-172)

